# A Prenylated dsRNA Sensor Protects Against Severe COVID-19 and is Absent in Horseshoe Bats

**DOI:** 10.1101/2021.05.05.21256681

**Authors:** Arthur Wickenhagen, Elena Sugrue, Spyros Lytras, Srikeerthana Kuchi, Matthew L Turnbull, Colin Loney, Vanessa Herder, Jay Allan, Innes Jarmson, Natalia Cameron-Ruiz, Margus Varjak, Rute M Pinto, Douglas G Stewart, Simon Swingler, Marko Noerenberg, Edward J D Greenwood, Thomas W M Crozier, Quan Gu, Sara Clohisey, Bo Wang, Fabio Trindade Maranhão Costa, Monique Freire Santana, Luiz Carlos de Lima Ferreira, ISARIC4C investigators, Joao Luiz Da Silva Filho, Matthias Marti, Richard J Stanton, Eddie C Y Wang, Alfredo Castello-Palomares, Antonia Ho, Kenneth Baillie, Ruth F Jarrett, David L Robertson, Massimo Palmarini, Paul J Lehner, Suzannah J Rihn, Sam J Wilson

**Affiliations:** MRC–University of Glasgow Centre for Virus Research, Institute of Infection, Inflammation and Immunity, University of Glasgow, UK; Cambridge Institute of Therapeutic Immunology and Infectious Disease, Jeffrey Cheah Biomedical Centre, Cambridge Biomedical Campus, University of Cambridge, Cambridge, CB2 0AW, UK; Roslin Institute, University of Edinburgh, United Kingdom; Laboratory of Tropical Diseases – Prof. Luiz Jacintho da Silva, Department of Genetics, Evolution, Microbiology and Immunology, Institute of Biology, University of Campinas, Campinas, SP, Brazil; Department of Education and Research, Oncology Control Centre of Amazonas State – FCECON, Manaus, AM, Brazil; Tropical Medicine Foundation Dr. Heitor Vieira Dourado, Manaus, AM, Brazil; Department of Pathology and Forensic Medicine, Tropical Medicine Foundation Dr. Heitor Vieira Dourado, Manaus, AM, Brazil; ISARIC4C investigators; Wellcome Centre for Molecular Parasitology, Institute of Infection, Immunity and Inflammation, University of Glasgow, UK; Division of Infection & Immunity, Cardiff University, Cardiff, United Kingdom

## Abstract

Cell autonomous antiviral defenses can inhibit the replication of viruses and reduce transmission and disease severity. To better understand the antiviral response to SARS-CoV-2, we used interferon-stimulated gene (ISG) expression screening to reveal that OAS1, through RNase L, potently inhibits SARS-CoV-2. We show that while some people can express a prenylated OAS1 variant, that is membrane-associated and blocks SARS-CoV-2 infection, other people express a cytosolic, nonprenylated OAS1 variant which does not detect SARS-CoV-2 (determined by the splice-acceptor SNP Rs10774671). Alleles encoding nonprenylated OAS1 predominate except in people of African descent. Importantly, in hospitalized patients, expression of prenylated OAS1 was associated with protection from severe COVID-19, suggesting this antiviral defense is a major component of a protective antiviral response. Remarkably, approximately 55 million years ago, retrotransposition ablated the OAS1 prenylation signal in horseshoe bats (the presumed source of SARS-CoV-2). Thus, SARS-CoV-2 never had to adapt to evade this defense. As prenylated OAS1 is widespread in animals, the billions of people that lack a prenylated OAS1 could make humans particularly vulnerable to the spillover of coronaviruses from horseshoe bats.

## Introduction and Results

Severe acute respiratory syndrome coronavirus 2 (SARS-CoV-2), which causes COVID-19, emerged in human populations in 2019 and has left an indelible mark on global health, culture and prosperity. SARS-CoV-2 is particularly sensitive to inhibition by type I interferons (IFNs) and there is great interest in understanding how individual IFN-stimulated genes (ISGs) inhibit SARS-CoV-2, as type I IFNs play a major role in governing COVID-19 disease outcome. Specifically, inborn errors (Zhang et al., 2020) and single nucleotide polymorphisms (Pairo-Castineira et al., 2020) within the IFN system are linked with more severe COVID-19. Moreover, neutralizing anti-IFN autoantibodies likely prevent host IFN responses from controlling SARS-CoV-2 replication, again resulting in severe COVID-19 disease (Bastard et al., 2020). Accordingly, recombinant IFNs have therapeutic potential (Monk et al., 2021) although the timing of IFN responses or the administration of recombinant IFNs is likely critical (Channappanavar et al., 2016).

We sought to better understand the protective role that IFNs play in COVID-19. We hypothesized that allelic and spliced variation in individual antiviral defenses might underlie the differential susceptibility to severe COVID-19 observed in different individuals. To identify the ISG products that inhibit SARS-CoV-2, we used arrayed ISG expression screening to identify effectors with anti-SARS-CoV-2 potential (Kane et al., 2016; Schoggins et al., 2011). We first confirmed that SARS-CoV-2 was IFN-sensitive in transformed human lung A549 cells modified to express the SARS-CoV-2 receptor ACE2 and the protease TMPRSS2 (Hoffmann et al., 2020; Rihn et al., 2021). These cells supported the efficient replication of SARS-CoV-2 and IFN pre-treatment attenuated multiple variants (including B.1.1.7) in an endpoint CPE assay (well-clearance (Rihn et al., 2021)) (Figure 1A). Thus, A549 cells were deemed suitable for ISG expression screens.

**Figure 1.**
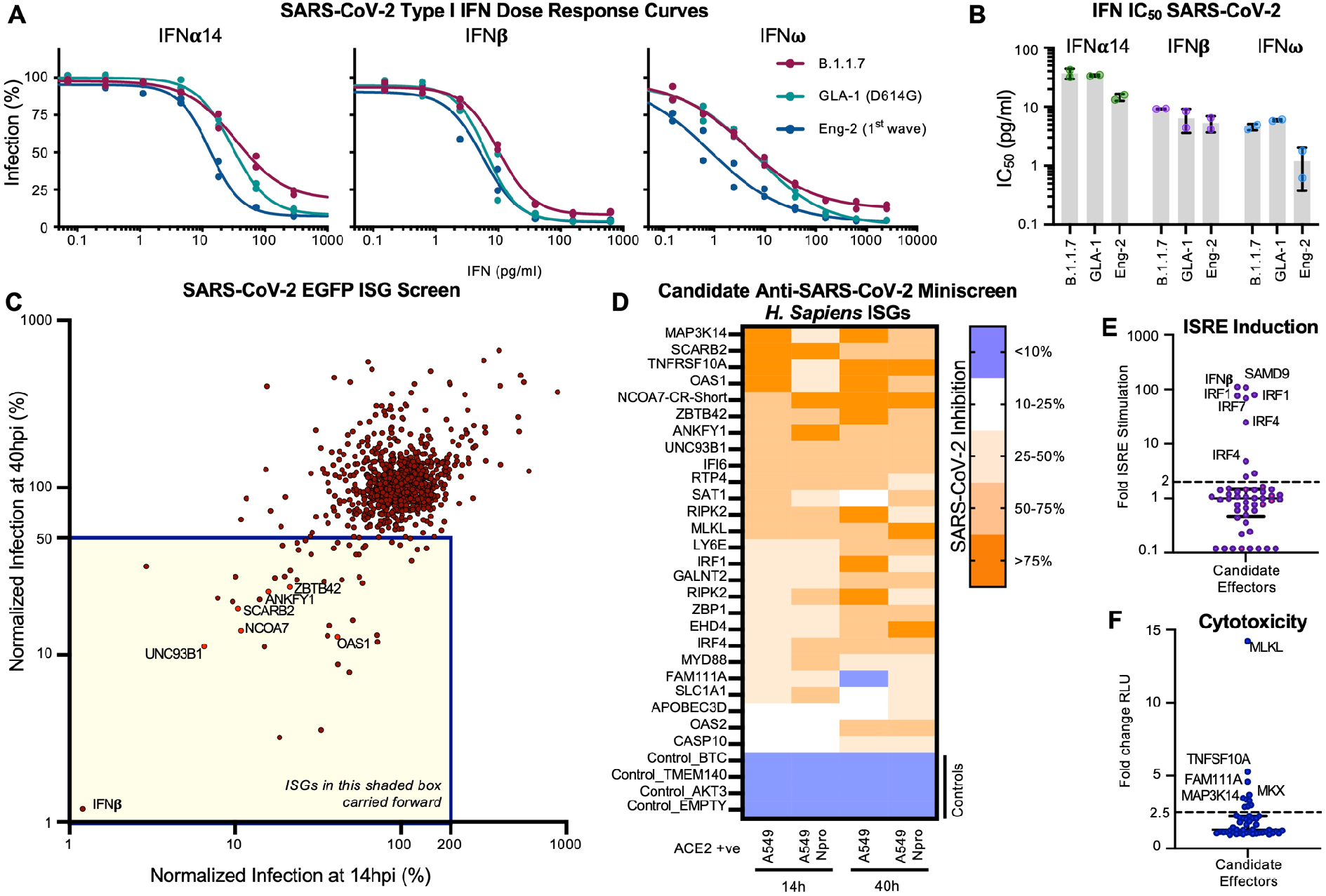
Arrayed ISG expression screening reveals factors with candidate anti-SARS-CoV-2 activity. (A) Dose response curves evaluating the ability of type I IFN pretreatment to protect A549 ACE2 TMPRSS2 cells from SARS-CoV-2 infection. Normalized infection (% well-clearance quantified by transmitted light). (B) IC_50_ estimates derived from (A). (C) A549 Npro cells were transduced with hundreds of individual human or macaque ISGs (see Supplementary Figure 1A-C) and infected with SARSCOV-2-EGFP (Wuhan-1), in duplicate, and the level of infection in the presence of each ISG was measured using flow cytometry at 14 and 40 hpi. (D) Miniscreen of the ability of human candidate effectors identified in (C) alongside controls, to inhibit SARSCoV-2 in A549 and A549 Npro at 14 and 40 hpi (the equivalent panel for macaque ISGs presented in Supplementary Figure 1J). (E,F) The ability of each human and macaque effector to stimulate ISRE activity using A549-ISRE-EGFP cells (E) or cause toxicity (Cyto Tox-Glo) (F) 48 hours posttransduction with the relevant ISG-encoding lentiviral vector. The dashed line indicates threshold for negative selection.

Because exogenously expressed ISGs can trigger antiviral gene expression programs (Kane et al., 2016), we selected transformed ACE2-expressing, IRF3-deficient A549 cells (A549 Npro ACE2), which possess an attenuated ability to produce IFN (Hale et al., 2009) as the background for the screens. We transduced these cells with an arrayed library of lentiviral vector-encoded ISGs in a 96-well plate format (one ISG per well), using a library of >500 human ISGs and a similar library of >350 rhesus macaque ISGs (Kane et al., 2016)(supplementary Figure 1A-C). The macaque ISGs were included as they increased the total number of ISGs under consideration. Importantly, ∼2/3 of the ISGs examined could potentially be relevant to betacoronavirus infection (Parkinson et al., 2020)(supplementary Figure 1D,E). In order to capture inhibition that might occur at any stage of the virus lifecycle, we used a GFP-encoding recombinant SARS-CoV-2 (Thi Nhu Thao et al., 2020) and measured the ability of each individual ISG to inhibit SARS-CoV-2 at 14 hours (early) and 40 hours (late) postinfection. Using this approach, we identified a number of candidate anti-SARS-CoV-2 effectors (Figure 1C). All candidates that conferred >2-fold inhibition at early and late timepoints or only at late timepoints were considered further using a series of independent confirmatory ‘miniscreens’. The magnitude of protection conferred by each ISG at early and late timepoints was assessed using ACE2-positive cells in the presence or absence of IRF3 (Figure 1D and Supplementary Figure 1F-J). In addition, we sought to subtract nonspecific inhibitors of SARS-CoV-2 by identifying ISGs that triggered a polygenic ‘antiviral state’ (Figure 1E) or caused cytotoxicity (Figure 1F). Following these confirmatory and negative selection screens, we identified six candidate antiviral effectors that robustly inhibited SARS-CoV-2 without inducing substantial toxicity or inducing ISRE expression.

The candidate effectors included known antiviral genes such as the IFN-inducible short isoform (isoform 4) of NCOA7, which inhibits influenza A viruses (IAVs) (Doyle et al., 2018), and OAS1, a dsRNA sensor capable of activating RNase L (Donovan et al., 2013; Han et al., 2014). We also identified UNC93B1, a polytopic membrane protein required for TLR trafficking (Kim et al., 2008; Majer et al., 2019) as well as SCARB2, a virus receptor (Yamayoshi et al., 2009) involved in cholesterol transport (Heybrock et al., 2019; Neculai et al., 2013). In addition, we identified ANKFY1 and ZBTB42, which have not previously been ascribed antiviral activity. We exogenously expressed these ISGs in human A549 lung cells modified to express ACE2 or modified to express both TMPRSS2 and ACE2 and examined their ability to inhibit a 1^st^ wave isolate of SARS-CoV-2 (CVR-GLA-1 (Rihn et al., 2021)). Interestingly, in the absence of TMPRSS2, all six candidate antiviral effectors inhibited the 1^st^ wave isolate to varying degrees (2.5-fold- >1000-fold)(Figure 2A), whereas only OAS1 consistently inhibited SARS-CoV-2 in both the presence and absence of TMPRSS2 (Figure 2A,B). Indeed, TMPRSS2-mediated entry has been proposed as a strategy used by SARS-CoV-2 to evade antiviral factors (Peacock et al., 2021).

**Figure 2.**
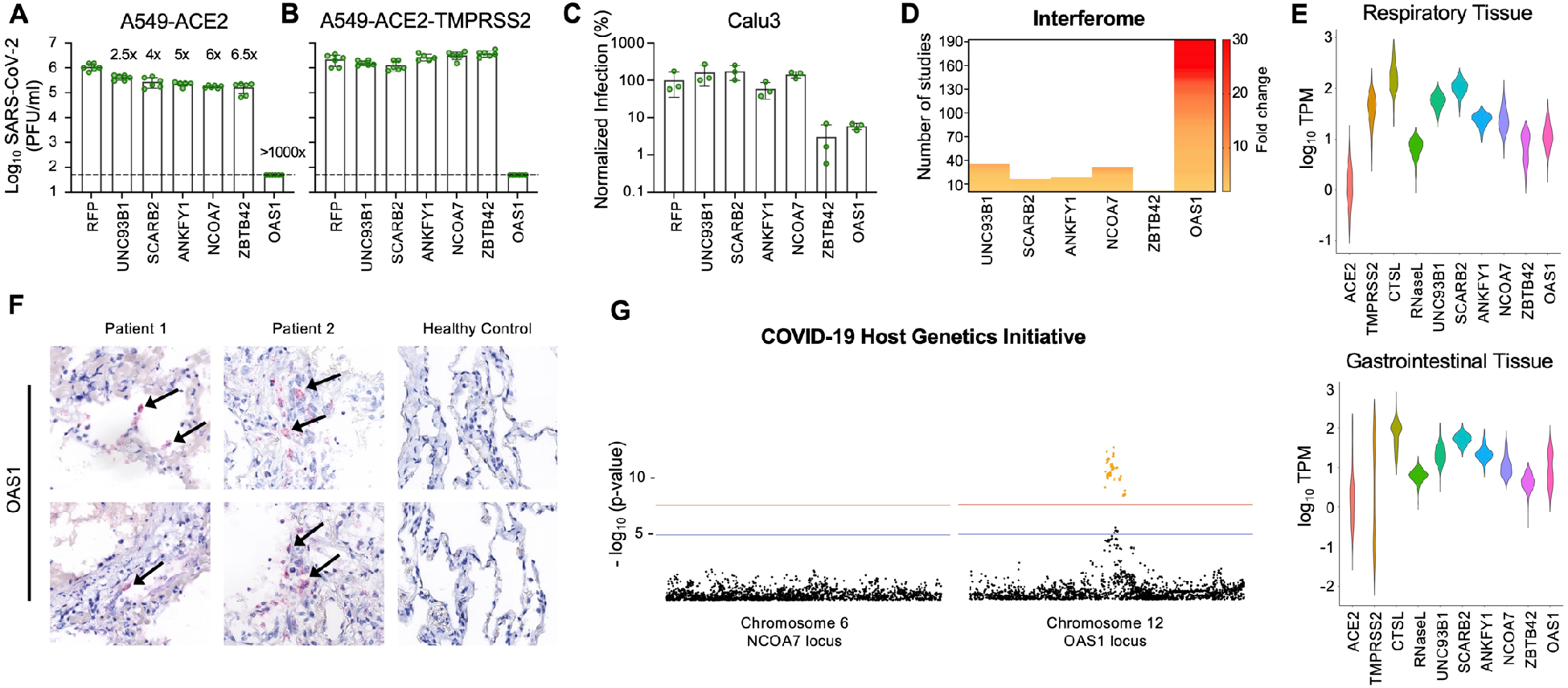
The ISG OAS1 strongly inhibits SARS-CoV-2. SARS-CoV-2 isolate GLA1 infectious titers (PFU) were determined using A549ACE2 (A) or A549-ACE2-TMPRSS2 cells (B) modified to express the ISG hits (UNC93B1, SCARB2, ANKFY1, NCOA7, ZBTB42 or OAS1) from the screening pipeline (Fig. 1 C-F). Fold-protection from SARS-CoV-2 is indicated for each gene in (A). (C) SARS-CoV-2-ZsGreen infectious titers on Calu3 cells expressing the same hit ISGs as in (A,B) measured by flow cytometry at 40 hpi. (D) ISG-ness of selected genes was assessed by fold-change upon type 1 IFN stimulation as reported in studies in the interferome database (interferome.org). (E) Gene expression analysis across different respiratory and gastrointestinal tissues using datasets from the Genotype-Tissue Expression (GTEx) database. ACE2 and TMPRSS2 included for reference. RNase L included as functionally linked to OAS1. (F) Detection of OAS1 gene expression by RNAscope in FFPE lung tissue of deceased COVID-19 patients compared to healthy control lung tissue. Arrows indicate staining +ve cells. (G) Meta-analysis of the COVID-19 Host Genetics Initiative (covid 19hg.org) for genetic variation between critical ill COVID19 patients and control populations at the gene locus of the NCOA7 and OAS1 genes with the red line indicating the threshold for significant SNPs (yellow dots).

To identify effectors present at the sites of SARS-CoV-2 infection, we examined the interferon responsiveness of the 6 candidate effectors from published studies using the interferome database (Figure 2D) (Rusinova et al., 2013), analyzed their basal expression in respiratory and gastrointestinal tracts (GTEx) (figure 2E) and assessed their transcript abundance in post-mortem lung tissue from COVID-19 patients (Figure 1F and Supplementary Figure 2). In addition, we examined the genomic locus of each candidate effector for SNPs that may be associated with increased susceptibility to infection and/or severe disease (Figure 2G and Supplementary Figure 2). Following these analyses, we focused our attention on OAS1 as the antiviral activity was the most robust (Figure 2A-C), basal OAS1 transcription is highly IFN inducible (Figure 2D,E), the mRNA is readily detectable in infected patients (Figure 2F) and the locus encompasses multiple SNPs that are associated with altered susceptibility to infection and severe disease (Figure 2G).

The canonical model of OAS antiviral activity involves initial dsRNA sensing by OASs, which results in the synthesis of 2′-5′-linked oligoadenylates (2-5A). 2-5A induces the dimerization of inactive RNase L, which then mediates the indiscriminate cleavage of viral and host RNAs presenting ssRNA UpU and UpA motifs (Bisbal and Silverman, 2007). The initial sensing of virus dsRNA that subsequently activates RNase L has mostly been ascribed to OAS3, with OAS1 infrequently considered as a major viral dsRNA sensor (Li et al., 2016). Indeed, in over 50 arrayed ISG screens completed in our laboratory, SARS-CoV-2 is the only virus substantially inhibited by OAS1 (Figure 3A). Surprisingly, while OAS1 potently inhibited SARS-CoV-2, exogenous OAS3 had no effect on the replication of SARS-CoV-2 (Figure 3B,C). To confirm that OAS1 was inhibiting SARS-CoV-2 through the synthesis of 2-5A and activated RNase L, we used CRISPR Cas9 to disrupt the RNase L locus in our modified permissive cells. The antiviral activity of OAS1 was only effective in the presence of RNase L and loss of RNase L abrogated the ability of OAS1 to inhibit SARS-CoV-2 (Figure 3D). RNase L activation can, in principle, inhibit viruses by degrading viral or host RNAs (Burke et al., 2019; Rath et al., 2019) (eventually resulting in apoptosis (Zhou, 1997)) or by triggering an IFN-response (Malathi et al., 2007). We examined the contribution of an RNase L-induced IFN response to the inhibition of SARS-CoV-2 by OAS1 by ablating JAK-STAT signaling using the Janus kinase (JAK) inhibitor ruxolintinib (Rux) (Quintás-Cardama et al., 2010). Type I IFN treatment potently inhibited SARS-CoV-2 replication (Figure 3E), and this effect was entirely reversed by the addition of Rux. Importantly, OAS1 potently inhibited SARS-CoV-2 in the absence of JAK-STAT signaling (Figure 3E) indicating that the RNase L-mediated destruction of host and/or viral RNAs is likely the predominant mechanism through which OAS1 inhibits SARS-CoV-2. Furthermore, in the absence of OAS1, SARS-CoV-2 replicated more effectively in the presence of Rux, suggesting that host IFN responses were still measurably inhibitory despite the antagonism strategies deployed by SARS-CoV-2 (Figure 3E).

**Figure 3.**
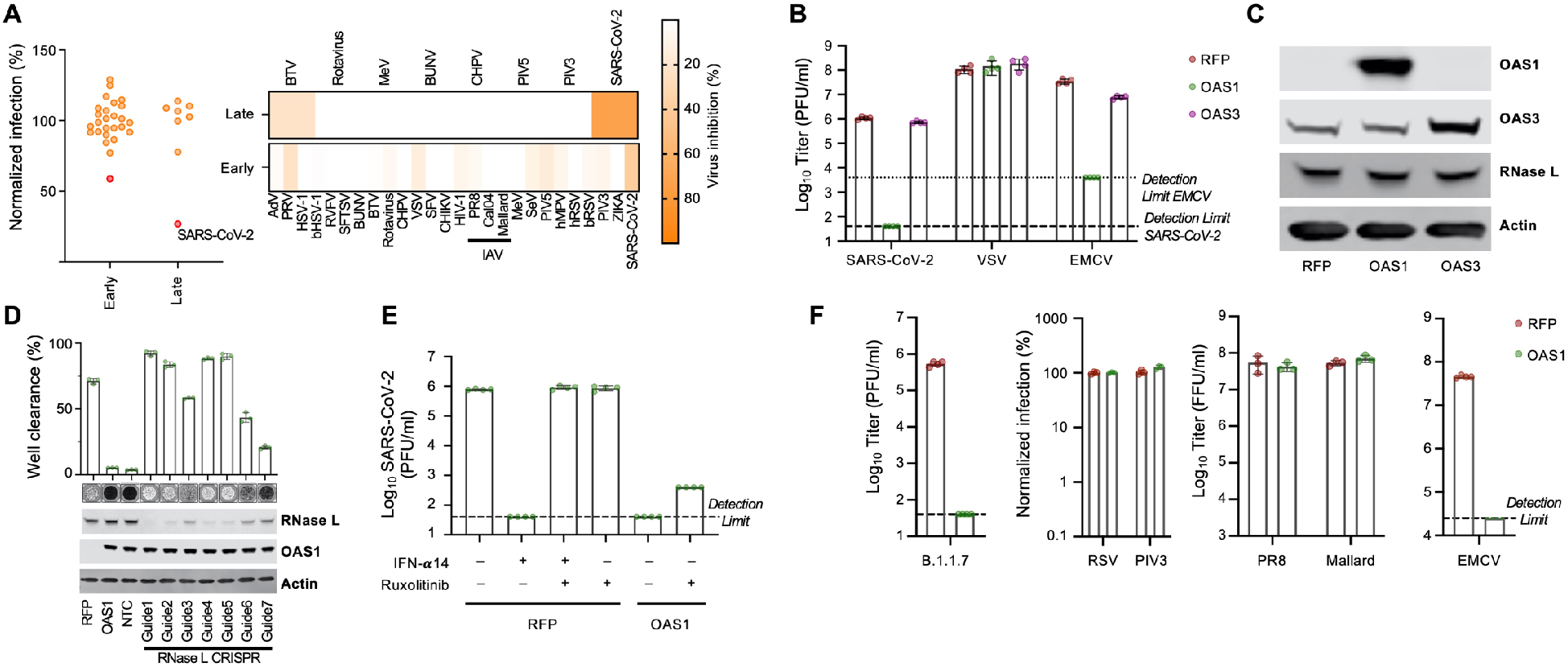
OAS1 inhibition of SARS-CoV-2 is specific and mediated via the RNase L pathway. (A) Normalized infection in the presence of OAS1 at early or late stages of the viral life cycle, quantified in large-scale ISG expression screens (as seen in Figure 1) for a panel of viruses. (B) A549ACE2-TMPRSS2 (AAT) cells were modified to express OAS1 and OAS3 and the titers of SARS-CoV-2, VSV and EMCV were determined (PFU) in the presence of each ISG. (C) Protein expression (OAS1, OAS3 and RNase L) in the cell lines tested in (B) was monitored using western blotting. (D) SARS-CoV-2 replication (well clearance at 72 hpi due to cytopathic effects of virus replication) was assessed in cells whose RNase L expression was reduced using seven different lentiviral vector-derived CRIPSR guides and one non-targeting control guide (NTC). The level of RNase L KO was assessed by Western blotting. (E) SARS-CoV-2 infectious titre (PFU) on AAT cells expressing RFP or OAS1 was determined in the presence and absence of pretreatment with 100U/ml IFNa 14 and/or 0.5 um Ruxolitinib. (F) AAT cells were modified to express RFP (control) or OAS1. The infectious titers of RSV-GFP and PIV3-GFP were determined using flow cytometry (24 hpi). Similarly, titers of influenza A viruses (PR8 and Mallard/H1N1) were determined using an immunostained focus forming assay and titers of SARS-CoV-2 B.1.1.7, EMCV and VSV were determined by plaque assay.

To better-understand why SARS-CoV-2 was inhibited by OAS1, we revisited the antiviral specificity of OAS1 by considering the ability of OAS1 to inhibit a panel of viruses that replicate via dsRNA intermediates within different subcellular compartments. We first confirmed that OAS1 was active against the more transmissible human-adapted B.1.1.7 variant, which remained highly sensitive to OAS1 restriction (Figure 3F). Intriguingly, when we examined three negative sense ssRNA viruses whose genome replication occurred in the cytosol (Indiana vesiculovirus (VSV), human respirovirus 3 (PIV-3) and human respiratory syncytial virus (RSV)), all were unaffected by OAS1 (Figure 3B,F).

Similarly, influenza A viruses (possessing a segmented negative sense ssRNA genome) whose replication occurs in the nucleus, were completely resistant to OAS1 (Figure 3F). In contrast, when we examined Cardiovirus A (EMCV), a positive sense ssRNA virus whose genome replication occurs within replicative organelles (ROs) and double-membrane vesicles (DMVs) (Melia et al., 2018), OAS1 restricted this virus by >100-fold (Figure 3B,F) (Chebath et al., 1987). Because CoVs replicate in similar ER-derived membranous structures to EMCV (Emerman et al., 2008; Romero-Brey and Bartenschlager, 2016; Snijder et al., 2006), we considered whether OAS1 might be a dsRNA sensor specifically targeted to membrane-bound ROs in infected cells.

In humans, OAS1 protein is expressed as two major forms designated p46 and p42. The longer p46 isoform (present in the screening library and used in Figures 1-3) is generated by alternative splicing to an exon downstream of the terminal exon utilized by the p42 isoform (Figure 4A). Although all human genotypes possess the exon that completes the transcript encoding p46, an intronic SNP (Rs10774671 aka 12-112919388-G-A) determines OAS1 exon usage. Individuals with a G variant of this SNP express the p46 isoform and some p42 whereas individuals with the A variant predominantly express the p42 isoform (and cannot express the p46 isoform) (Bonnevie-Nielsen et al., 2005; Li et al., 2017; Noguchi et al., 2013). Individuals with G at this SNP are more resistant to West Nile virus infection (Lim et al., 2009) and respond better to IFN-therapy following hepatitis C infection (El Awady et al., 2011). They are also less susceptible to multiple sclerosis (O’Brien et al., 2010) and Sjögren’s syndrome (Li et al., 2017) which may involve virus infection. Importantly, this SNP was also associated with protection against severe COVID-19 disease (Pairo-Castineira et al., 2020; Zeberg and Pääbo, 2021; Zhou et al., 2021). We thus determined whether p42 possessed the same anti-SARS-CoV-2 activity as the p46. Remarkably, the p42 isoform, which is the most common isoform in humans (∼61%) possessed no detectable anti-SARS-CoV-2 activity (Figure 4B). Although differential basal enzymatic activity has been proposed to underlie the divergent antiviral potential of p46 and p42 OAS1 (Bonnevie-Nielsen et al., 2005), this effect is likely due to expression level as p42 and p46 possess similar catalytic activity (Carey et al., 2019). Remarkably, the C-terminus of p46 encodes a canonical CAAX-box prenylation signal (CTIL) that is absent from the p42 variant (Figure 4C), and predicted to be geranylgeranylated (Maurer-Stroh and Eisenhaber, 2005). Indeed, prenylation of OAS1 was proposed to alter the subcellular localization of OAS1, perhaps influencing mitochondrial respiration (Skrivergaard et al., 2019). We hypothesized that prenylated OAS1 is targeted to membranous viral replicative organelles and facilitates the sensing of CoV dsRNA (and perhaps many divergent RNA viruses that replicate within replicative organelles). Remarkably, introducing a point mutation into the p46 isoform, that prevented its prenylation (C397A), completely ablated the antiviral activity of p46 (Figure 4B). Similarly, appending a four amino acid CAAX-box (CTIL) to the C-terminus of the p42 isoform conferred substantial antiviral activity to the inactive p42 form, reducing the ability of SARS-CoV-2 to form plaques by more than 100-fold (Figure 4B). Thus, prenylation of OAS1 appears necessary for dsRNA sensing of SARS-CoV-2. A nearly identical picture emerged using EMCV (Figure 4D). While prenylated p46 and p42-CTIL reduced EMCV plaque formation by >100-fold, nonprenylated p42 or p46 C397A possessed no anti-EMCV activity. Importantly, this antiviral activity of p46 and p42-CTIL was highly specific and did not inhibit the ability of VSV to form plaques in parallel experiments (Figure 4E).

**Figure 4.**
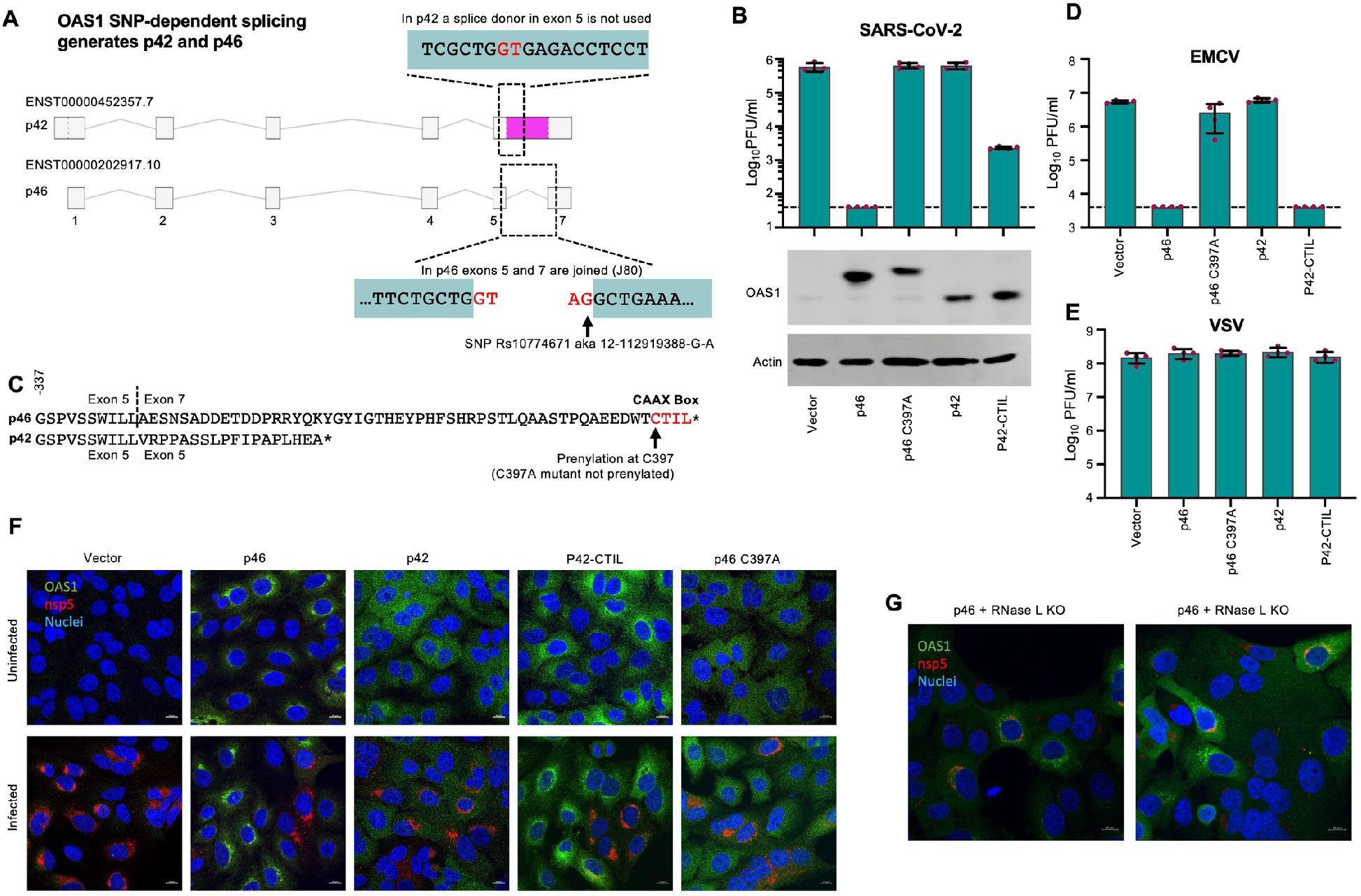
OAS1 isoforms have differential antiviral activity as determined by C-terminal prenylation. (A) Schematic representation of OAS1 splicing resulting in isoforms p42 and p46. (B) SARS-CoV-2 infectious titer on AAT cells expressing the OAS1 isoforms p46, not prenylated p46 (p46 C397A), P42 or prenylated p42 (P42-CTIL) or a vector control. Protein expression analysis of the levels of isoforms and mutants is shown by Western blot. (C) Protein sequence alignment of the p42 and p46 isoforms indicating the CAAX-box prenylation signal in 246. (D) EMCV infectious titer and (E) VSV infectious titer on the cells from (B) as determined by plaque assay (PFU). (F) Immunofluorescence on cells from (B) infected with SARS-CoV-2 isolate GLA1 at MOI 0.5 for 24h followed by staining with anti-OAS1 (green) and anti-SARS-CoV2-nsp5 (red) antibodies and nuclear Hoechst stain (blue). Contrast was reduced in the p46 sample to prevent oversaturation in the green channel due to particularly strong perinuclear concentration. (G) As in (F) using p46 RNase L KO cells (Guide 1, LHS and Guide 5, RHS) from Figure 3D

Because SARS-CoV-2 replication, which uses dsRNA intermediates, occurs within membranous replicative organelles, we next considered whether prenylation localizes OAS1 to these replicative organelles. Prenylated p46 and p42-CTIL localized to membranous perinuclear structures reminiscent of the ER (Figure 4F), whereas nonprenylated p42 and p46 C397A were diffusely distributed (Figure 4F). The subcellular localization of OAS1 was largely dependent on its prenylation status as prenylated p42-CTIL showed the same endomembranous localization as p46, whereas nonprenylated p46 C397A was diffusely distributed. To determine whether prenylated OAS1 localized to SARS-CoV-2 replicative organelles, we co-stained infected cells with the viral nsp5 (Rihn et al., 2021). However, the p46 block was sufficiently strong to prevent formation of nsp5+ replicative structures, which were only visible in clusters of cells expressing low levels of p46 (Figure 4F). To overcome this, we imaged infected OAS1 expressing cells, whose RNase L expression was decreased using CRISPR Cas9 (Figure 4G). Relieving the block to SARS-CoV-2 replication imposed by OAS1, showed colocalization of OAS1 with the SARS-CoV-2 replicative organelles (Figure 4G).

The realization that prenylation is essential for OAS1-mediated sensing of SARS-CoV-2 allowed us to examine the transcriptome of infected patients and investigate whether there is a link between the expression of prenylated OAS1 and SARS-CoV-2 disease progression. The multiple OAS1 alleles encode 4 common p46 protein variants (Figure 5A), all of conferred protection against SARS-CoV-2 infection (Figure 5B). Because each p46 variant possessed antiviral activity (i.e. was not confined to a single haplotype), we examined the p46 splice junction directly (Figure 4A) to assess whether expression of p46-encoding mRNA, as opposed to the presence or absence of SNPs (Pairo-Castineira et al., 2020), significantly influenced the severity of COVID-19. Importantly, the frequency of the Rs10774671 G SNP that governs the expression of p46 varies between ∼11% (Peruvian in Lima) and ∼70% (Esan in Nigeria) (Figure 5C) and could have a major influence on the susceptibility of different populations to severe COVID-19.

**Figure 5.**
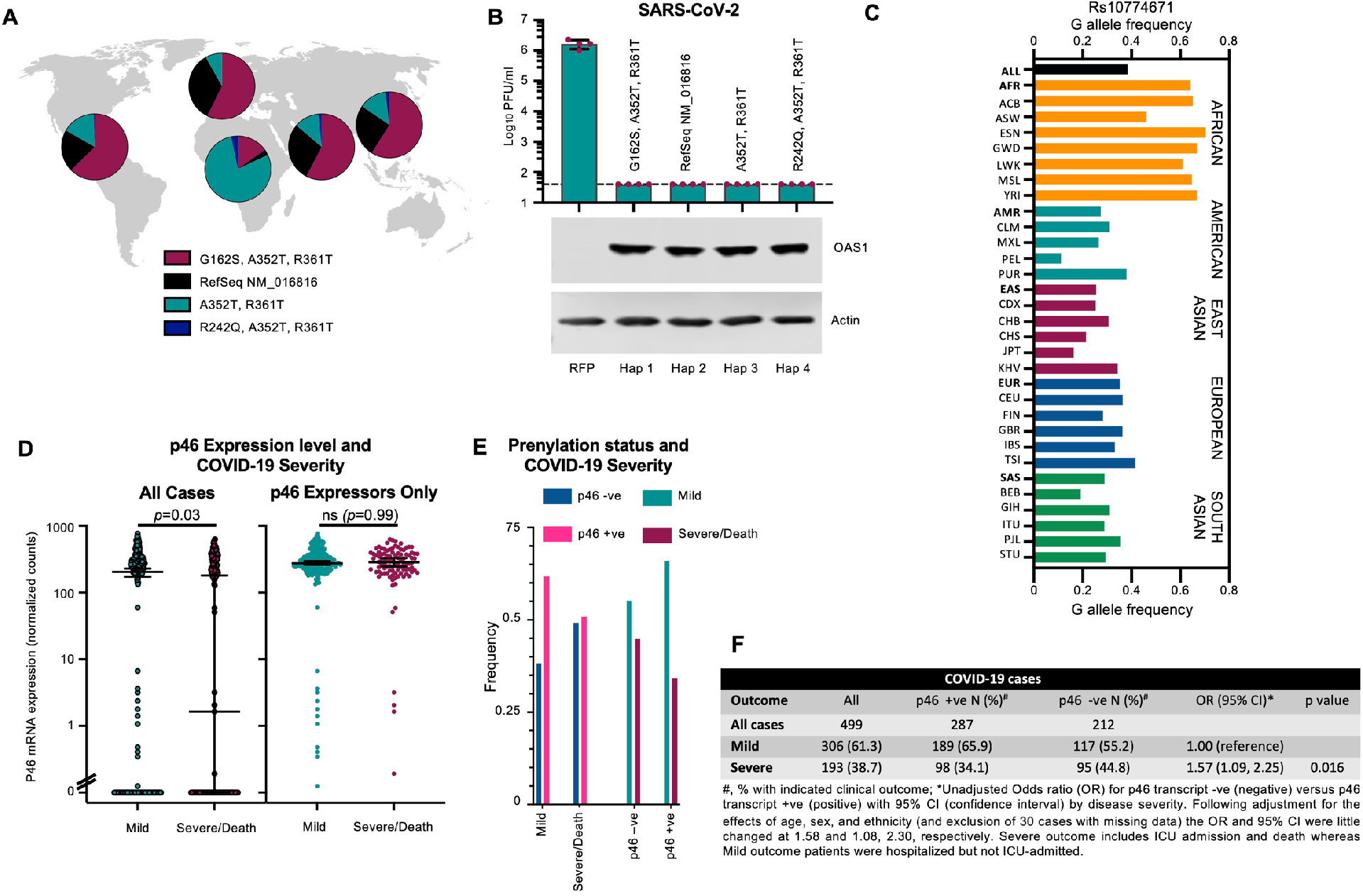
Prenylated OAS1 protects against severe COVID-19. (A) Allelic frequencies of the most common circulating p46 variants of OAS1. (B) Infectious titers of SARS-CoV-2 CVR GLA-1 (PFU) were determined on AAT cells modified to express OAS1 of each human p46 variant. OAS1 expression was monitored using western blotting (lower panels). (C) Frequency of alleles with G at Rs 10774671 in different human populations (1000 genomes project). (D) Transcript abundance of the p46 isoform (encoding prenylated OAS1), determined using Junction Seq analysis of RNAseq data from PBMCs from infected patients with mild (hospitalized but not ICU-admitted) or severe/lethal (ICU-admitted and/or death) COVID19. Significance was determined using a Mann-Whitney U test. (E) Prenylation status (p46 -ve or +ve), determined by the presence or absence of P46 transcript (from D) in mild and severe COVID-19. (F) Tabulated odds ratios and 95% confidence intervals of the data presented in D and E.

Although the p46 transcript encodes 54 C-terminal amino acids (aa) that are not part of the p42 protein (Figure 4C), individuals homozygous for A at Rs10774671 (AA) can form splice junctions 1 nucleotide downstream of the p46 splice junction. We therefore confirmed that we could reliably detect the absence of p46 in RNAseq data derived from AA cells. We used JunctionSeq to examine all OAS1 transcript junctions (annotated on Ensembl) in AA cells in the presence or absence of IFN treatment (Supplementary Figure 4). Accordingly, we were unable to detect the specific junction (J80) between exons 5 and 7 that specifies the expression of prenylated OAS1 in AA cells (Supplementary Figure 4).

We therefore applied this method to RNAseq data from 499 hospitalized UK COVID-19 patients with known disease outcomes (ISARIC4C). We defined severe outcomes as intensive care unit (ICU) admission and/or death whereas mild outcome patients were hospitalized but not ICU-admitted. All patients expressed OAS1 and we detected a substantial number of individuals who expressed OAS1 p42 but did not express detectable OAS1 p46 (Supplementary Figure 5). Specifically, 212/499 individuals (42.5%) did not express p46

Remarkably, the absence of prenylated OAS1 was associated with more severe disease (Figure 5E-F). Specifically, the median transcript abundance of p46 was over 100-fold lower in the severe COVID-19 group (Figure 5D). This difference was entirely driven by the over-representation of patients that did not express any prenylated OAS1 in the severe COVID-19 group (Figure 5E). p46 mRNA levels were almost identical in individuals that expressed prenylated OAS1, regardless of whether they experienced mild or severe COVID-19 (Figure 5D). Thus, patients lacking the p46 transcript were significantly more likely to have severe disease (95/212, 44.8%) compared to those expressing p46 (98/287, 34.1%) (unadjusted Odds Ratio (OR)=1.57, 95% CI 1.09, 2.25; following adjustment for age, sex, and ethnicity and exclusion of 30 cases with missing data OR=1.58, 95% CI 1.08, 2.30) (Figure 5F). Death was also more frequent in these patients (34/212, 16% versus 34/287, 11.8%) with the effect size similar to that for disease severity, but these differences were not statistically significant (unadjusted OR 1.42, 95% CI 0.85, 2.37). Taken together, our analysis of OAS1 antiviral activity *in vitro*, combined with our analysis of OAS1 transcripts in patient cohorts, reveals that the protective Neanderthal OAS1 haplotype (Zeberg and Pääbo, 2021; Zhou et al., 2021) prevents severe disease through specifying the expression of prenylated OAS1, which directs dsRNA sensing to the sites of SARS-CoV-2 replication.

The specific antiviral defenses in a given species can form a barrier to the cross-species transmission of viruses. As the differential splicing of p46 and p42 isoforms is poorly characterized beyond primates, it has been difficult to investigate the protection conferred by p46 in non-human species. The realization that prenylation underlies the differential antiviral activity allowed us to investigate this aspect of OAS1 beyond humans. For example, many coronaviruses encode phosphodiesterases (PDEs) that degrade 2-5A and antagonize OAS protein activity (Zhao et al., 2012). The human betacoronavirus OC43 encodes such a phosphodiesterase (NS2) and as predicted (Goldstein et al., 2017), OAS1 failed to inhibit OC43 (Figure 6A). OC43 likely entered human populations via a cross-species transmission from cows (Vijgen et al., 2005), and subsequent examination of the bovine genome identified 3 OAS1 paralogues, one of which is prenylated (OAS1Y). In accordance with Figure 4, only the prenylated bovine OAS1Y conferred potent anti-SARS-CoV-2 activity (Figure 6B, note that we were unable to confirm efficient expression of OAS1X using polyclonal antibody raised to human OAS1). Thus, the extreme sensitivity of SARS-CoV-2 to prenylated OAS1 and the absence of OAS1 antagonism in SARS-CoV and SARS-CoV-2 led us to determine whether horseshoe bats possess a prenylated OAS1 defense. The OAS1 locus in the greater horseshoe bat (*Rhinolophus ferrumequinum*) does not contain a transcript or exon encoding a prenylated OAS1. Indeed, all Ensembl /NCBI database entries specified nonprenylated proteins. Remarkably, when we examined the genomic region where the prenylation signal should reside (based on synteny and homologous flanking sequences), retrotransposition of an LTR sequence was seen, ablating the CAAX-box motif, and therefore preventing the expression of prenylated anti-CoV OAS1 in these bats (Figure 6C). We searched for this insertion in 44 available bat genome sequences and identified the same insertion only in members of the *Rhinolophoidea* superfamily (including *Rhinolophus, Hipposideros* and *Megaderma* species), indicating that this ancient retrotransposition insertion occurred ∼58-52 million years ago (MYA), within this bat superfamily. In contrast, we could detect CAAX-box encoding syntenic sequences in members of all other bat taxa (Figure 6D). Importantly, this means that membrane-associated, prenylated, p46-like, OAS1 dsRNA sensing has been ablated in the bat reservoirs of SARS-CoV and SARS-CoV-2 and suggests why the horseshoe bats might be such prolific reservoir hosts of Betacoronaviruses. The absence of pressure to avoid prenylated OAS1 in these bats may have allowed SARS-CoV-2 to be particularly sensitive to this pattern recognition pathway.

**Figure 6.**
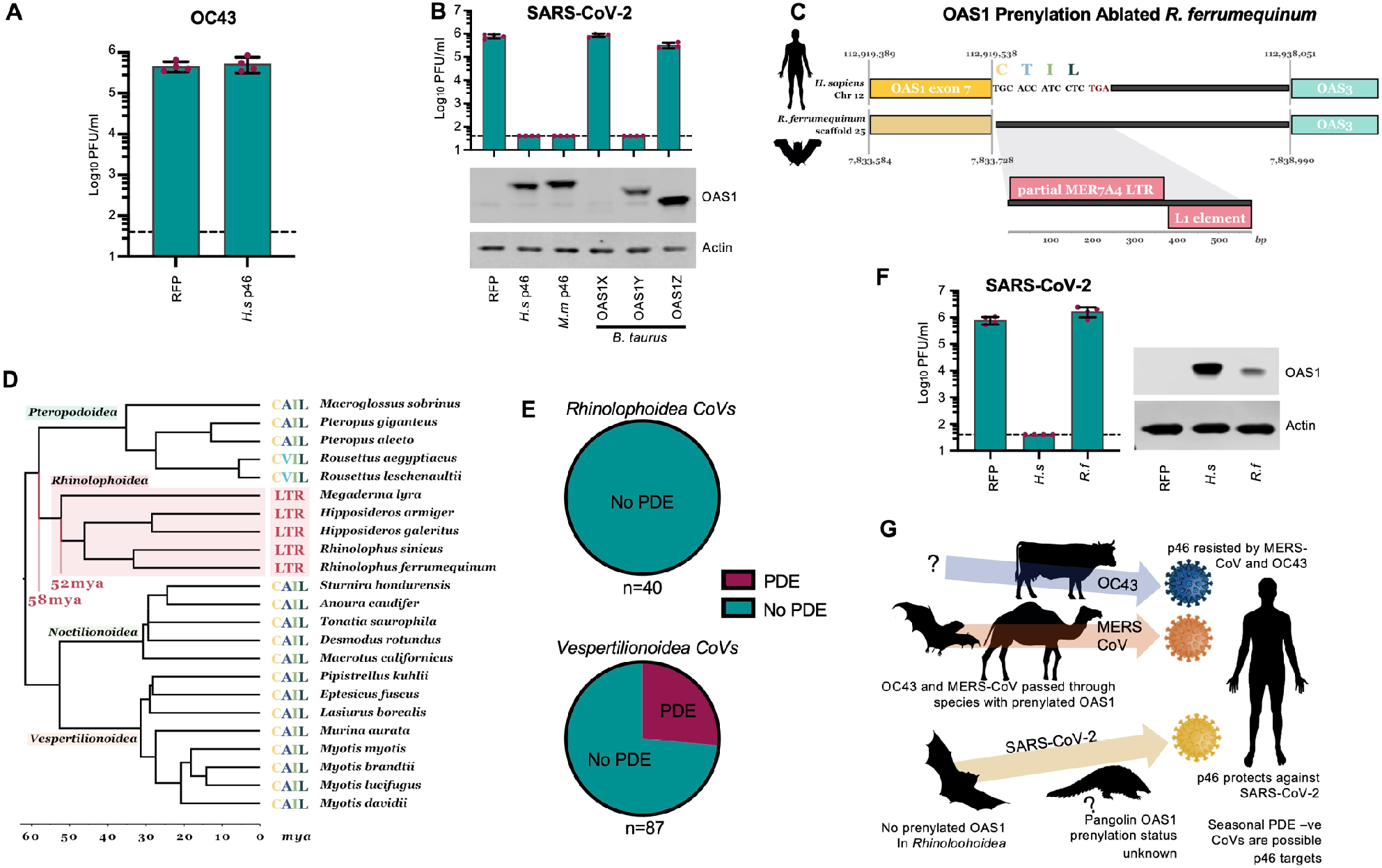
Retrotransposition at the OAS1 locus in *Rhinolophoidea* has ablated the sequence that encoded a CAAX-box prenylation signal. (A) Infectious titers of OC43 (PFU) were determined on AAT cells modified to express OAS1 from humans (H.S p46). (B) Infectious titers of SARS-CoV-2 CVR GLA-1 (PFU) were determined on AAT cells modified to express OAS1 from humans (H.s), macaques, (*M.m*) and cows (B. taurus). OAS1 expression was monitored by western blotting (lower panels) (C) Schematic of genome synteny between the human OAS1 exon 7 locus (yellow) and the *R. ferrumequinum* genome. The exact syntenic sequence coordinates are annotated for the start of OAS1 exon 7, the start of the CAAX-box encoding sequence and the start of the upstream gene locus, OAS3 (blue). Transposable element hits on the 580bp non-syntenic region in the *R. ferrumequinum* genome are shown in the zoomed in inset. Non-coding regions are shown in black. Note that the schematic is not to scale. (D) Dated phylogeny (retrieved from timetree; www.timetree.org) of bat species with a confirmed LTR insertion in the OAS1 locus or a CAAX-box encoding sequence present in the same scaffold as their OAS1 locus. Clades are labelled by superfamily, species names and CAAX sequence (or LTR) are annotated next to the tree tips. The approximate time period during which the LTR insertion took place is annotated in red.(E) Pie charts of CoVs from *Rhinolophoidea* and *Vespertilionoidea* binned according to whether they are known or predicted to encode a phosphodiesterase (PDE) OAS antagonist. (F) Infectious titers of SARS-CoV-2 CVR GLA-1 (PFU) were determined on AAT cells modified to express OAS1 from humans (*H.s*) and horseshoe bats (R.f). OAS1 expression was monitored by western blotting (right-hand panels) (G) A diagrammatic representation highlighting that the OAS1 prenylation status in reservoir and intermediate species likely determines whether a CoV is sensitive or resistant to p46-mediated activation of Rnase L. Cow prenylated OAS1 (AY243505), dromedary camel prenylated OAS1 (KAB1255732.1), pipistrellus prenylated OAS1 (XM 036409709).

Considering the lack of prenylation in *Rhinolophoidea* OAS1 and the ability of coronavirus PDEs to antagonise this pattern recognition pathway, we sought to examine whether PDE-encoding coronaviruses infect horseshoe bats. Given the variability in coronavirus encoded PDEs (NS4b in Lineage C and NS2 in Lineage A Betacoronaviruses (Goldstein et al., 2017; Thornbrough et al., 2016)), we developed a custom HMM protein profile using NS4b, NS2, the mammalian PDE AKAP7 (Gusho et al., 2014) and rotavirus A VP3 (Zhang et al., 2013) PDE sequences screened through all putative open reading frames (ORFs) of all published *Coronaviridae* genomes. This method should capture previously unannotated or undescribed PDEs. Although the available sequence data set is likely biased by sampling, no known *Rhinolophoidea* infecting coronaviruses encode PDEs. In fact, all bat viruses with PDEs detected were collected from bats in the *Vespertilionoidea* superfamily (whose prenylated OAS1 is intact).

Based on the absence of PDEs in SARS-CoV-2 and the lack of prenylated OAS1 in *Rhinolophoidea*, we predicted the OAS1 transcript of *Rhinolophus ferrumequinum* to be inactive against SARS-CoV-2. Accordingly, OAS1 from greater horseshoe bats did not inhibit SARS-CoV-2 in our assays (Figure 6F). We speculate that the absence of pressure to evade prenylated OAS1 (in horseshoe bats) has left SARS-CoV-2 particularly sensitive to this defense (when encountered in human populations). In contrast, CoVs that originate from species with prenylated OAS1 (Figure 6G) are likely to be less susceptible to this prenylated dsRNA sensor (Figure 6G).

## Discussion

Extant species are selected to adapt to their environment. Viruses are no exception and even SARS-CoV-2, a ‘generalist’ virus (MacLean et al., 2021), has likely adapted to replicate in the animal reservoir(s) in which it circulates. Cross-species transmission to humans exposed SARS-CoV-2 to a new repertoire of antiviral defenses, some of which SARS-CoV-2 may not have been selected to evade. Prenylated OAS1 may be an example of such a defense with prenylated OAS1 being targeted to endomembranous structures where it exhibits potent anti-SARS-CoV-2 activity *in vitro*.

Importantly, hospitalized COVID-19 patients lacking the p46 transcript had worse clinical outcomes compared to those who expressed prenylated OAS1. Severe disease was significantly more frequent with ICU admission or death being approximately 1.6 times more likely in these patients. The increased risk associated with the p42 phenotype is similar to the increased risk for male sex among the same hospitalized patients analysed in this study (data not shown). The increased odds of death was similarly raised among patients lacking p46. However, the number of deaths in this cohort was relatively small and the differences observed were not statistically significant. A larger study is needed to provide enough power to determine whether lack of p46 transcript is also associated with increased mortality. We could not detect an association between disease severity and p46 transcript abundance in individuals who express p46. This could reflect the fact that expression level is less important for an enzyme than a saturable restriction factor (because enzyme catalysis greatly amplifies pattern recognition). Alternatively, p46 expression in whole blood may not reflect expression differences at the sites of viral replication or differences in p46 expression level occurring earlier in infection (before hospitalization). Importantly, geographical variation in the frequency of alleles encoding prenylated OAS1 could contribute to predisposing some populations, such as people of African descent, to be more resistant to severe COVID-19.

OAS1 is part of the evolutionary ‘core’ interferon response, meaning that OAS1 has been maintained as interferon responsive for over 300 million years (Shaw et al., 2017). Indeed, prenylated forms of OAS appear to be evolutionarily ancient and can be found in nearly all mammals and even in divergent vertebrates (such as XM_033043060 in fish). One troubling consequence of the allelic variation in OAS1 expression is that only a minority of individuals are homozygous for alleles that encode prenylated OAS1. In fact, humans may be relatively unusual mammals, as billions of humans are predicted to be unable to express prenylated OAS1 at all. For example, in Chinese populations, the G variant is present at a frequency of ∼21-31% (depending on the region). Accordingly, ∼48-62% of individuals are homozygous for A and unable to express prenylated OAS1, and fewer than 10% are homozygous for the G variant encoding prenylated OAS1. It is therefore possible that because large numbers of humans lack this defense, humans are especially vulnerable to the cross-species transmission of sarbecoviruses from horseshoe bats. More work is needed to assess the OAS1 locus, and OAS1 allelic diversity, in animal species known to support the replication and spread of SARS-like viruses such as palm civets, raccoon dogs, pangolins and mink, to investigate whether prenylated OAS1 might form a barrier to the initial cross-species transmission of sarbecoviruses. Although there is a paucity of genome sequence data for these species, we have thus far been unable to identify transcripts encoding prenylated OAS1 in raccoon dogs, palm civets or pangolins.

There is currently great interest in identifying the biological characteristics of bats that might predispose them to be reservoirs of circulating viruses and much work has focused on innate immunity (Banerjee et al., 2020). However, it is important to be cautious when generalizing about bats as all species possess unique innate immune features (Shaw et al., 2017). More importantly, bats are an extraordinarily diverse order (>1300 species) and individual bat species may not be any more likely to act as viral reservoirs than other species (Mollentze and Streicker, 2020). Nonetheless, it is striking that horseshoe bats not only lack prenylated OAS1-mediated dsRNA sensing, but have a reduced ability to sense cytosolic DNA through STING (Xie et al., 2018). Thus it is tempting to speculate that multiple defects in pathogen recognition makes horseshoe bats particularly good virus reservoirs.

The generation of cytosolic and membrane-associated OAS1 through alternative splicing is not solely a human strategy and multipIe primate species can express p42 and p46 OAS1 isoforms (Carey et al., 2019). Thus, encoding prenylated OAS1 is likely the ancestral state and humans have gained and/or maintained alleles that cannot express p46. In light of the apparent inactivity of OAS1 p42, it is notable that the p42-encoding alleles predominate and are more common in all human populations (apart from people of African descent). Accordingly, the p46 variant may have been selected against, possibly because it is deleterious in the absence of virus infection (Carey et al., 2019; Field et al., 2005). Accordingly, a catalytically inactive OAS1 has been identified in tamarin monkeys (Carey et al., 2019), suggesting that complete loss of OAS1 may have been beneficial to this species. However, the repeated convergent evolution of nonprenylated OAS1 variants, through independent mechanisms including alternative splicing (Bonnevie-Nielsen et al., 2005), premature stop codons (Carey et al., 2019) or retention of expanded paralogues (Perelygin et al., 2006), suggests that nonprenylated OAS1 has been positively selected, perhaps to sense viral pathogens that replicate in the cytosol. If p42-encoding OAS1 alleles were previously advantageous to humans, this trend has recently reversed as the p46-encoding Neanderthal-like OAS1 haplotype has increased in frequency over the last millennium in Eurasia perhaps driven by historic pandemics (Zeberg and Pääbo, 2021).

Although directing OAS1 dsRNA sensing to CoV ROs via C-terminal prenylation, followed by activation of RNase L is the mechanism through which OAS1 inhibits SARS-CoV-2, some questions remain. Specifically, whether RNase L inhibits SARS-CoV-2 directly, through degrading viral RNAs, or whether inhibition is indirect through degrading host mRNAs is currently unknown. Moreover, the observation that many CoVs invest part of their genomic potential in coding for PDEs, suggests that CoVs tend to present attractive PAMPs for recognition by OASs. Whether CoVs present specific motifs that are sensed by OAS1 or whether targeting to the ROs is sufficient to facilitate relatively nonspecific recognition of viral dsRNA will require further work to resolve. An additional implication of this study is that the protective effect of prenylated OAS1 against Sjögren’s syndrome (Li et al., 2017) and multiple sclerosis (O’Brien et al., 2010), suggests that viruses that replicate in association with endomembranous structures could be potential triggers for these disorders.

Thus far, expressing prenylated OAS1 has prevented severe COVID-19 in a substantial fraction of infected individuals (likely measured in ‘hundreds of thousands’ in the UK alone). With the continued emergence of SARS-CoV-2 variants of concern, it is reassuring that the B.1.1.7 variant retains susceptibility to detection by prenylated OAS1. However, as the pandemic continues, and new lineages arise, it will be important to remain vigilant. For example, the recombinant acquisition of a PDE gene (from a seasonal CoV or host gene) or selection against specific dsRNA motifs recognized by prenylated OAS1 could increase the pathogenicity of SARS-CoV-2, reinforcing the need to pay close attention to the phenotypic properties of emerging SARS-CoV-2 variants.

## Materials and Methods

### Cell lines, plasmids and viruses

All cells were maintained in Dulbecco’s modified Eagle’s medium (DMEM) supplemented with 10% FCS and 10 μg/ml gentamicin unless otherwise stated. A549-ACE2-TMPRSS2 (‘AAT’) and VeroE6-ACE2-TMPRSS2 (‘VAT’) cells have been described previously (Rihn et al., 2021). HEK-293T cells were propagated from lab stocks and A549-NPro cells were a kind gift of Prof. Richard E. Randall. Calu-3 cells were maintained in MEM supplemented with 10% FCS, 2 mM glutamine, 2 mM sodium pyruvate and 100 μM non-essential amino acids.

The SARS-CoV-2 viruses CVR-GLA-1 and SARS-CoV-2-ZsGreen have been described previously (Rihn et al., 2021). SynSARS-CoV-2-eGFP was a kind gift from Prof. Volker Thiel (Thi Nhu Thao et al., 2020). B.1.1.7 was isolated from a clinical sample (kind gift of Prof. Wendy Barclay). Indiana vesiculovirus (VSV) was a kind gift of Dr. Megan Stanifer and was propagated on BHK cells at low MOI as described previously (Rihn et al., 2019). Influenza A viruses A/Puerto Rico/8/1934 (H1N1) and A/mallard/Netherlands/10-Cam/1999(H1N1) were rescued from reverse genetics systems (a kind gift from Prof. Ron Fouchier, and Prof. Laurence Tiley, respectively) as described previously (de Wit et al., 2004; Turnbull et al., 2016). Human respirovirus 3 with GFP (PIV3-GFP) was purchased from ViraTree. Human respiratory syncytial virus expressing GFP (RSV-GFP) was a kind gift from Prof. Peter Collins (Hallak et al., 2000). Encephalomyocarditis virus (EMCV) was a kind gift from Dr. Connor Bamford. Betacoronavirus OC43 (ATCC VR-1558) was purchased from ATCC and propagated on VAT cells.

### Retroviral vectors and cell modification

The lentiviral vector pSCRPSY (KT368137.1) has been previously described (Kane et al., 2016). pLV-EF1a-IRES-Puro (Addgene plasmid # 85132) was modified by PCR amplifying the tagRFP ORF (using pSCRPSY as template) flanked by directional SfiI sites which were further flanked by BamHI and EcoRI restriction sites (forward oligo: 5’-CTC TCG GAT CCG GCC GAG AGG GCC ATG AGC GAG CTG ATT AAG-3’ and reverse oligo: 5’-CTC TCG AAT TCG GCC AGA GAG GCC TCA CTT GTG CCC CAG-3’) and the BamHI-EcoRI fragment was subcloned into the vector to create the modified pLV-EF1a-IRES-Puro-SfiI-tagRFP construct. The cDNA corresponding to the ORFs of the following OAS genes (GenBank accession number): OAS1p46 (NM_016816), human OAS3 (NM_006187), macaque OAS1 (EF467665), bovine OAS1X (NM_178108), bovine OAS1Y (NM_001040606), bovine OAS1Z (AY650038), and Rhinolophus ferrumequinum OAS1 (XM_033097132 / Ensembl: ENSRFET00010016745) were chemically synthesised as gene-blocks with flanking SfiI sites (IDT DNA) and the SfiI fragment was subcloned into the modified pLV-EF1a-IRES-Puro-SfiI plasmid. To generate the human OAS1p42 sequence (in accordance with GenBank accession NM_002534), OAS1p46-c397a, and OAS1p42-CTIL sequences, the pLV-SfiI-OAS1p46 lentiviral vector plasmid was modified by overlap extension PCR (using primer pair 5’-CTC TCT GGC CGA GAG GGC CAT GAT GGA TCT CAG AAA TAC CCC AG-3’ and 5’-TCT CTC GGC CAG AGA GGC CTC AAG CTT CAT GGA GAG GGG CAG GGA TGA ATG GCA GGG AGG AAG CAG GAG GTC TCA CCA GCA GAA TCC AGG AGC TCA CTG GG-3’ for OAS1p42, primer pair 5’-CTC TCT GGC CGA GAG GGC CAT GAT GGA TCT CAG AAA TAC CCC AG-3’ and 5’-TCT CTC GGC CAG AGA GGC CTC AGA GGA TGG TGG CGG TCC AGT CCT CTT CTG CCT GTG GG-3’ for p46-c395a, and primer pair 5’-CTC TCT GGC CGA GAG GGC CAT GAT GGA TCT CAG AAA TAC CCC AG -3’ and 5’-TCT CTC GGC CAG AGA GGC CTC AGA GGA TGG TGC AAG CTT CAT GGA GAG GGG CAG GGA TGA ATG GCA GGG AGG AAG CAG GAG GTC TCA CCA GCA GAA TCC AGG AGC TC ACT GGG -3’ for p42-CTIL) and the SfiI-fragment was subcloned in place of OAS1p46 in the pLV lentiviral vector plasmid described above. Lentiviral vectors were produced by transfecting 293T cells as described previously (Rihn et al., 2019) and 0.45 μm-pore-size-filtered supernatant was used to transduce AAT cells and were selected using 2 μg/ml puromycin.

Gene editing by CRISPR/Cas9 was achieved using the lentiCRISPRv2-BlastR one vector system (Sanjana et al., 2014; Shalem et al., 2014) following the established protocols from the Zhang laboratory. CRISPR guides were designed using the CHOPCHOP online tool (https://chopchop.cbu.uib.no). Seven guides were sub-cloned into the one vector system between the BsmBI sites using annealed oligonucleotides with directional, compatible BsmBI overhangs and tested in their efficacy to ablate RNase L expression (guide 1: 5’-CAC CGC CGA GTT GCT GTG CAA ACG-3’, guide 2: 5’-CAC CGT TAT CCT CGC AGC GAT TGC G-3’, guide 3: 5’-CAC CGC TAT AGG ACG CTT CGG AAT G -3’, guide 4 5’-CAC CGT ATA GGA CGC TTC GGA ATG T-3’, guide 5: 5’-CAC CGT AGT CAT CTT CAG CCG CTA T-3’, guide 6: 5’-CAC CGT TTA TCC TCG CAG CGA TTG C-3’, and guide 7: 5’-CAC CGC AAT CGC TGC GAG GAT AAA -3’) and a non-targeting control guide (‘NTC’: 5’-GTG ACG TAC CGC TGG AGG TA-3’) was used as a control. Transduced cells were selected and cultured in medium additionally supplemented with 5 μg/ml blasticidin (Melford Laboratories).

### Arrayed ISG expression screening

The ISG overexpression libraries and flow cytometry based screening have been described previously (Feng et al., 2018; Kane et al., 2016; Schoggins et al., 2011). Briefly, two lentiviral vector ISG libraries consisting of 539 human and 444 macaque ISGs were used to transduce A549-NPro-ACE2 cells in the presence of polybrene for 48 h, allowing ISG expression from an early HIV-1 mRNA and tagRFP from an unspliced late HIV-1 mRNA, the latter used as a marker for transduction. Transduced cells were then infected with synSARS-CoV-2-eGFP (Thi Nhu Thao et al., 2020) in the presence of DMEM supplemented with 2% FCS. At 14 h or 40 h post-infection, cells were trypsinised and fixed in 4% formaldehyde. Percentage of transduced cells (tagRFP positive) and SARS-CoV-2 infected cells (GFP-positive) were determined by flow cytometry using a Guava EasyCyte flow cytometer (Millipore).

A549-ISRE::GFP cells (gift from Prof. Richard E.Randall, (Stewart et al., 2014)) were transduced in the presence of polybrene with the ‘miniscreen’ library of selected ISGs from the SARS-CoV-2 screening. 96 h post-transduction the supernatant was harvested to measure toxicity of the expressed ISGs using the CytoTox-Glow kit (Promega) and cells were fixed in 4% formaldehyde to measure ISRE induction (GFP-positive cells) as surrogate for IFN induction.

### Virus infections and titrations

SARS-CoV-2 infection assays, plaque assays and well-clearance assays have been described previously (Rihn et al., 2021). Briefly, well-clearance assays quantify transmitted light (Celigo, Nexcelom) through imaging of a stained cell monolayer. CPE induced clearance of the monolayer transmits more light compared to uninfected or protected monolayers. Virus inputs were normalized using plaque assays on VeroE6 cells to 500 PFU per well. For interferon and ruxolitinib treatment, cells were pre-treated with doses specified in text/figures for 2 h prior to infection, and the equivalent concentration was included in the culture medium post-infection. Plaque assays with VSV, EMCV and OC43 were performed under the same conditions as SARS-CoV-2 for 2 (VSV and EMCV) or 5 days (OC43).

For titration of GFP-expressing viruses (PIV3-GFP and RSV-GFP), 96-well plates of cells were infected with serial dilutions of virus for specified times and percentage GFP-positive cells was measured using flow cytometry with a Guava EasyCyte cytometer (Millipore). IAV foci immunostaining was achieved using mouse the anti-influenza A virus nucleoprotein monoclonal antibody clone AA5H (BioRad, MCA400) and visualised with goat anti-mouse IgG (H+L)-HRP conjugate (BioRad, 1721011) and TrueBlue Peroxidase Substrate (KPL, 5510-0030) following standard plaque assay protocols as described previously (Turnbull et al., 2016).

### Western blot analyses

For preparation of cell lysates, cells were washed once with PBS and then harvested in SDS sample buffer (12.5% glycerol, 175 mM Tris-HCl [pH 8.5], 2.5% SDS, 70 mM 2-mercaptoethanol, 0.5% bromophenol blue). Proteins were subsequently separated on NuPage 4% to 12% Bis-Tris polyacrylamide gels and transferred onto nitrocellulose membranes. Blots were probed with either antibodies raised against actin (mouse JLA20 hybridoma; courtesy of the Developmental Studies Hybridoma Bank, University of Iowa), OAS1 (rabbit polyclonal 14955-1-AP, Proteintech), OAS3 (rabbit polyclonal 21915-1-AP, Proteintech), or the rabbit anti-RNase L monoclonal antibody (Cell Signalling Technology, 27281). Thereafter, membranes were probed with species IgG-specific fluorescently labelled secondary antibodies goat anti-rabbit IgG or goat anti-mouse IgG (Thermo Scientific) and scanned using a LiCor Odyssey scanner.

### Immunofluorescence

Sub-confluent AAT derivative cells seeded on glass coverslips were infected with CVR-GLA-1 at an MOI of 0.5 for 24 h. Cells were fixed in PBS/8% formaldehyde, permeabilised with PBS/0.2% TX-100, and blocked with PBS/1% BSA. Immunostaining was performed using a rabbit anti-OAS1 monoclonal antibody [clone D1W3A] (Cell Signaling Technology, 14498) and sheep anti-SARS-CoV-2-nsp5 antiserum, (https://mrcppu-covid.bio described in (Rihn et al., 2021)). Secondary antibody staining was performed with Alexa Fluor™ 488 Goat anti-Rabbit IgG and Alexa Fluor™ 594 Donkey anti Sheep IgG both at 1:1000 (Invitrogen). Hoechst 33342 was included in the secondary antibody stain at 5 μg/ml. Coverslips were mounted on glass slides (VWR) using ProLong™ Gold antifade mountant (Thermo Fisher Scientific).

Maximum intensity projection (MIP) images of cell monolayers were acquired with an Airyscan Fast detector fitted to a Zeiss LSM880 confocal microscope. The objective lens used was a Plan-Apochromat 63x/1.4 oil DIC M27 (Carl Zeiss) and gain, laser power and pinhole were synchronised across images. MIP images with pixel scaling of 0.04µm x 0.04µm each comprised a Z stack of 10 individual slices with a total focal depth of 1.435µm. The OAS1-Alexa 488 was excited at 488 nm and detected in the 495-550 nm range, nsp5-Alexa 594 was excited at 594 nm and detected with a long pass 605nm filter and Hoechst was excited at 405 nm and detected in the 420-480 nm range. Post-acquisition, the contrast of images within each set were optimised using Zen software (Carl Zeiss) to equal degrees for the vector, p42, p42-CTIL and p46 C397A samples, whereas the histogram maximum was increased independently in the p46 sample shown in (Figure 4) to prevent oversaturation in the green channel due to strong perinuclear concentration. Images were acquired as 8-bit *.czi files and exported as 8-bit TIFF files.

### *In situ*-hybridisation

Formalin-fixed and paraffin-embedded (FFPE) lung tissue of 2 patients with confirmed SARS-CoV-2 infection (C21-20, C19-20) were used. As control tissue, FFPE lung of a healthy, 62-years old, male donor was used (NBP2-30182, Novusbio, cat. No 0028000B).

For the detection of Gene-specific RNA by *in situ*-hybridisation, the RNAscope 2.5 HD Reagent Kit-RED (code: 322350, Advanced Cell Diagnostics) and the probes (code: NCOA7 1029911-C1, ZBTB42 1029921-C1, OAS1 1029931-C1, ANKFY1 1029941-C1, UNC93B1 1029951-C1, SCARB2 1029961-C1; Advanced Cell Diagnostics) were designed (gene bank no: NM_001199622, NM_001137601, NM_016816, NM_016376, NM_030930, NM_005506) and purchased. As positive and negative control, a human ubiquitin and a plant probe were used, respectively (codes: 310041 and 310043, Advanced Cell Diagnostics). The protocol was followed according to the manufacturer’s instructions. This work (ethics approval number 32077020.6.0000.0005) was approved on May 2020 by the National Committee in Ethics and Research, Brazil. in COMISSÃO NACIONAL DE ÉTICA EM PESQUISA.

### Synteny analysis

The Ensembl web database (Yates et al., 2020) was used for assessing the OAS1 locus genome synteny between the human genome (GRCh38.p13) and the available *Rhinolophus* species, *R. ferrumequinum*, genome (mRhiFer1_v1.p). The syntenic region between the human OAS1 exon 7 (ENSE00003913305) and the horseshoe bat genome was examined to identify a region in the latter genome sequence starting at position 7,833,728 of scaffold 25 of the mRhiFer1_v1.p primary assembly that lacked synteny to the human genome. Incidentally, the non-syntenic region started in-frame where the p46 ‘CTIL’ encoding human sequence would have been (Supplementary Table 1).

We extracted the 580bp *R. ferrumequinum* sequence span up to where synteny resumes to the human genome and used hmmscan (HMMER 3.2.1) (Eddy, 2009) to search against the Dfam database (Hubley et al., 2015) for transposable elements present in the sequence. Two confident matches were identified, one to a partial MER74A-like LTR element at the very start of the non-syntenic sequence and one to a L1-like retrotransposon element at the 3’ end of the sequence (Supplementary Table 1).

### *In silico* genome screening

To explore how far back in time this LTR insertion at the OAS1 locus took place we used the Database-integrated genome-screening (DIGS) software (Zhu et al., 2018). DIGS uses a nucleotide or amino acid sequence probe to perform a BLAST similarity search through genome assemblies. We collected a set of 44 *Chiroptera* species genome assemblies (Supplementary Table 1). to perform three *in-silico* screens.

We first used the nucleotide sequence of the syntenic region of *R. ferrumequinum* to human exon 7 (Ensembl) and the adjacent 580bp region with the detected LTR insertion until homology resumes to the human genome as a probe (Supplementary digs_probes.fas). The DIGS screen was conducted using a minimum blastn bitscore of 30 and minimum sequence length of 30 nucleotides. Matches were aligned using MAFFT v7.453 (Katoh and Standley, 2013) and inspected for covering all regions of the probe (Supplementary Table 1). The second screen used the CAAX terminal amino acid sequence - homologous to that encoded by the human exon 7 - of 5 previously annotated bat OAS1 proteins holding a CAAX terminus (Supplementary digs_probes.fas). The minimum tblastn bitscore was set to 60 and minimum sequence length to 40 nucleotides. The translated sequences of hits were aligned and only hits with a CAAX domain present were retained (Supplementary Table 1). Finally, to cross-validate that sequence hits of the CAAX domain search are part of the OAS1 locus, we performed a screen using the *R. ferrumequinum* OAS1 C-terminal domain amino acid sequence as a probe (Supplementary digs_probes.fas) with a minimum tblastn bitscore of 100 and minimum sequence length of 100 nucleotides. Only hits of the CAAX search found on scaffolds with a detected OAS1 locus in the third search were maintained in the analysis. It is worth noting that lack of an OAS1 domain detected on the same scaffold as hits in the CAAX sequence search is most likely a result of low genome assembly quality. Regardless the hits were excluded for completeness.

### PDE analysis

To examine the diversity of PDE proteins encoded by coronaviruses we first constructed an HMMER protein profile. Two seemingly independently acquired PDEs are encoded by the NS2 of Lineage A Betacoronaviruses (Goldstein et al., 2017) and NS4b of MERS-like coronaviruses (Thornbrough et al., 2016) respectively. Group A rotavirus (RVA) has also been described to encode a protein with a homologous PDE domain and similar biological function (Zhang et al., 2013). Finally, the AKAP7 mammalian protein holds a PDE domain that has been experimentally shown to complement the function of murine coronaviruses’ NS2 activity (Gusho et al., 2014). We aligned the amino acid sequence of the PDE domains of the OC43 NS2 (AAT84352.1), the MERS NS4b (AIA22866.1) and the NS4b proteins of 2 more bat *Merbecoviruses* HKU5 (YP_001039965.1) and SC2013 (AHY61340.1), the AKAP7 proteins of *Rattus norvegicus* (NP_001001801.1), *Mus musculus* (NP_001366167.1) and humans (NP_057461.2) (as their homology to CoV PDEs has been previously characterised) and the Rotavirus A VP3 protein (AKD32168.1). The alignment was then manually curated using Bioedit based on the homology described in the literature. The final alignment was used to produce an HMM profile using the HMMER suite (v3.2.1) (Eddy, 2009).

All complete *Coronaviridae* sequences were downloaded from the NCBI Virus online database (Hatcher et al., 2016). Only sequences with an annotated host and length above 25,000 bp were retained and viruses of ‘Severe acute respiratory syndrome-related coronavirus’ species with a human host were excluded, producing a dataset of 2042 complete or near-complete coronavirus genomes. The EMBOSS getorf program (Rice et al., 2000) was used to extract the translated sequences of all methionine starting open reading frames (ORFs) with length above 100 nucleotides from the filtered virus genome dataset. All putative ORFs were then screened against our custom PDE HMM profile using hmmscan (Eddy, 2009).

### ISG expression in gastrointestinal and respiratory tissue

The 6 key genes (UNC93B1, SCARB2, ANKFY1, NCOA7, ZBTB42, OAS1) that were hits in our screens, SARS-CoV-2 cofactors (ACE2, TMPRSS2, CSTL) and RNase L were examined for their gene expression across respiratory and gastrointestinal tissues using 17,382 RNA-sequencing datasets from the Genotype-Tissue Expression (GTEx) v8 database (GTExConsortium, 2020). Gene expression is shown as log10 transform of transcripts per million (TPM). The respiratory tissue here includes Lung and Minor Salivary Gland tissues, while gastrointestinal tissue includes Colon, Esophagus, and Small Intestine tissues.

### MAIC analysis

The background dataset in MAIC analysis was created as described previously (Parkinson et al., 2020). MAIC was then run with the human and macaque lists, each independently, to determine the overlap between these lists and the manually curated systematic review of host factors associated with betacoronavirus literature.

### Clinical data analysis

499 whole-blood patient transcriptomes with known disease outcomes were obtained from the ISARIC4C consortium (https://isaric4c.net/). Ethical approval was given by the South Central-Oxford C Research Ethics Committee in England (reference 13/SC/0149), and by the Scotland A Research Ethics Committee (reference 20/SS/0028). The study was registered at https://www.isrctn.com/ISRCTN66726260.

Pre-processed and STAR (Dobin et al., 2013) mapped paired-end reads of 499 whole-blood patient transcriptomes with known disease outcomes from the ISARIC4C study were analysed to stratify mild (hospitalised but not ICU-admitted patients) and severe (ICU admitted and/or deceased) patients further into p46 +ve and p46 -ve groups. Using alignment files as input, strand-specific splice-junction-level counts for each sample were generated using QoRTs (Quality of RNA-seq Tool-Set) (Hartley and Mullikin, 2015). QoRTs generates a set of non-overlapping transcript features from the genome annotation, assigns a unique identifier to each feature, and generates counts for each annotated transcript subunit. Based on the presence or absence of p46 junction counts, mild and severe patient samples were sub-divided further into p46 +ve (Mild and SEV) and p46 -ve (Mild and SEV) groups. JunctionSeq (Hartley and Mullikin, 2016) was used to perform differential usage analysis of both exons and splice junctions, using a design model specifying sample/group types. Differential usage results of p46 junction (named J80 by the QoRTs) were interpreted and visualised using visualisation functions of JunctionSeq.

Disease severity and survival were compared in patients with and without expression of the p46 transcript. Severe disease was classified as ‘ICU admission’ or death and mild disease as ‘no ICU admission’ and alive at discharge from hospital. Survival was classified as death or ‘alive at discharge from hospital’. Binary logistic regression was used to estimate odds ratios (ORs) and 95% confidence intervals (CIs) with and without adjustment for the effects of age, sex and ethnicity. Analyses were implemented using IBM SPSS Statistics version 25 (IBM Corp. Armonk, USA). The G frequencies in different populations (at Rs10774671) were extracted from the 1000 genomes project (Sudmant et al., 2015).

## Data Availability

All data included or available upon final publication (unless prevented by ethical or legal constraints).

## Acknowledgements

This report is independent research which used data provided by the MRC funded ISARIC4C Consortium and which the Consortium collected under a research contract funded by the National Institute for Health Research. The views expressed in this publication are those of the author(s) and not necessarily those of the ISARIC4C consortium.

We thank Paul Moss, Peter Openshaw and the UK Coronavirus Immunology Consortium (UK CIC) for their guidance and suggestions and thank Volker Thiel, Wendy Barclay and the G2P-UK consortium for viruses. We also thank Afiq Aziz and Wil Furnon for technical assistance. Thanks to Nexcelom (Celigo) for their technical assistance and support during the UK COVID-19 lockdown (in 2020). The ISARIC WHO CCP-UK study protocol is available at http://isaric4c.net/protocols; study registry https://www.isrctn.com/ISRCTN66726260. This work uses data provided by patients and collected by the NHS as part of their care and support #DataSavesLives. We are grateful to the 2,648 frontline NHS clinical and research staff and volunteer medical students who collected the data in challenging circumstances and the generosity of the participants and their families for their individual contributions in these difficult times. We also acknowledge the support of Jeremy J. Farrar and Nahoko Shindo.

## Funding

This work was partly funded by UKRI/NIHR through the UK Coronavirus Immunology Consortium (UK-CIC MR/V028448/1 to MP and SJW) and the MRC via the following grants, MR/ K024752/1(to SJW), MC_UU_12014/10 (to MP and SJW) and MR/P022642/1 (to SJW and SJR), MR/V000489/1 (to ECYW and RJS), MR/S00971X/1 (to RJS and ECYW), MR/P001602/1 (to ECYW), MR/V011561/1 (to PJL). Support was also provided by a Wellcome Principal Research Fellowship 210688/Z/18/Z (to PJL), a Wellcome Trust Fellowship 201366/Z/16/Z (to SJR), the Addenbrooke’s Charitable Trust and the NIHR Cambridge Biomedical Research Centre (to PJL), support from the German Research Foundation, Deutsche Forschungsgemeinschaft; project number 406109949 (to VH), and German Federal Ministry of Food and Agriculture through BMEL Förderkennzeichen: 01KI1723G (to VH), and a Daphne Jackson Fellowship funded by Medical Research Scotland (to SS).

---

**Supplementary Figure 1.**
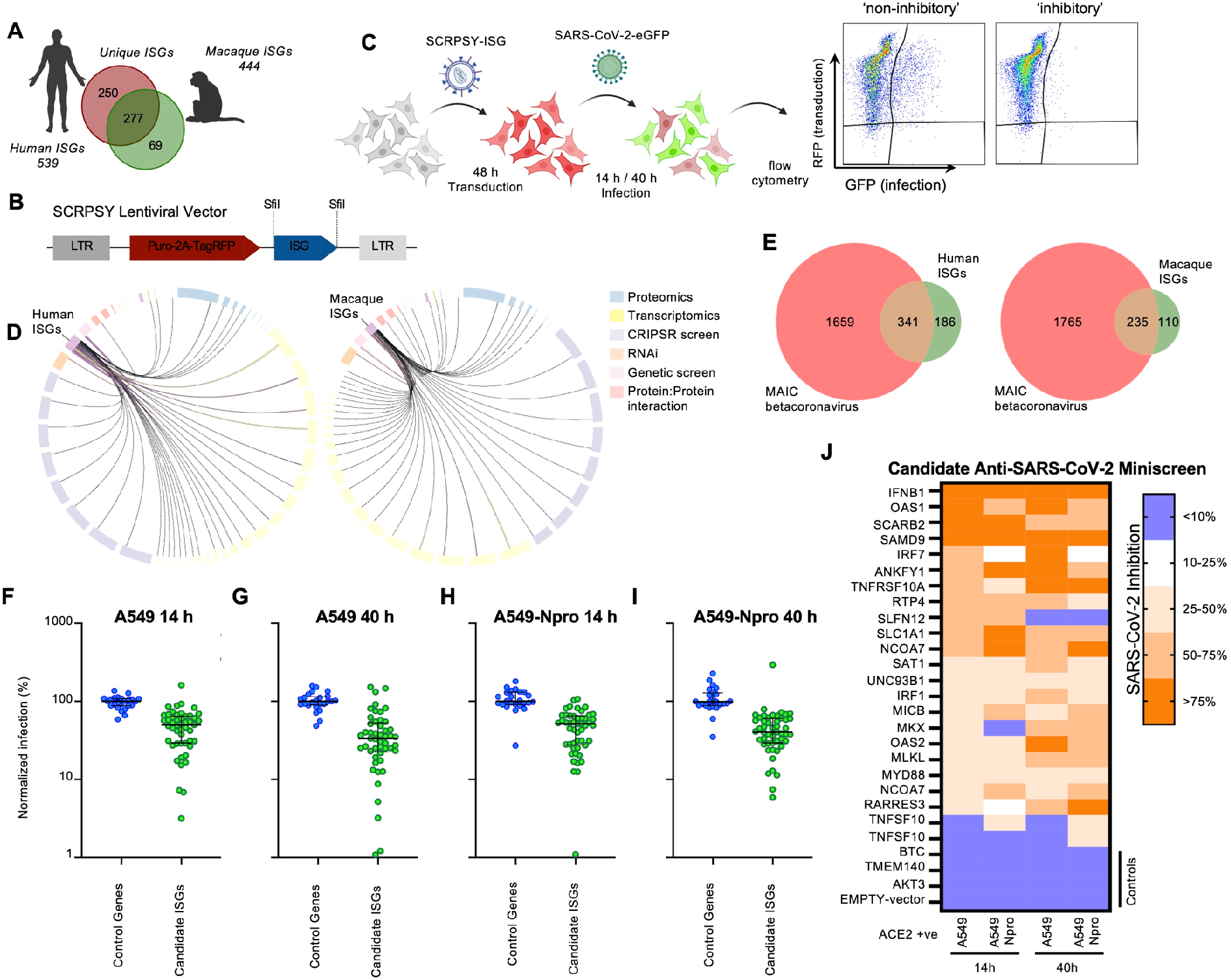
Arrayed ISG expression screening schematic and extended data. (A) A diagrammatic representation of the ISG libraries used. (B) A diagrammatic representation of the SCRPSY lentiviral vector used in (C, F-J) and in Figure 1C-F. (C) Schematic diagram of the ISG screening method used in Figure 1C and confirmatory ‘miniscreens’ (created with biorender.com). (D) Circos diagram illustrating the overlap between ISG candidates and datasets curated from betacoronavirus literature. ISG candidates are well supported with evidence from many experimental types. (E) Venn diagram of overlap between the Human ISG candidates (left) and Macaque ISG candidates (right) with the top 2000 hits from the betacoronavirus MAIC analysis (release: 25/11/2020). (F-1) Scatter plots of the individual miniscreens represented in (J) and Figure 1D on A549 (F-G) or A549-Npro (H-I) at 14 or 40 hpi. (J) Miniscreen of the ability of macaque candidate effectors identified in Figure 1C alongside controls, to inhibit SARSCOV-2 in A549 and A549 Npro at 14 and 40 hpi (the equivalent panel for human ISGs presented in Figure 1D).

**Supplementary Figure 2.**
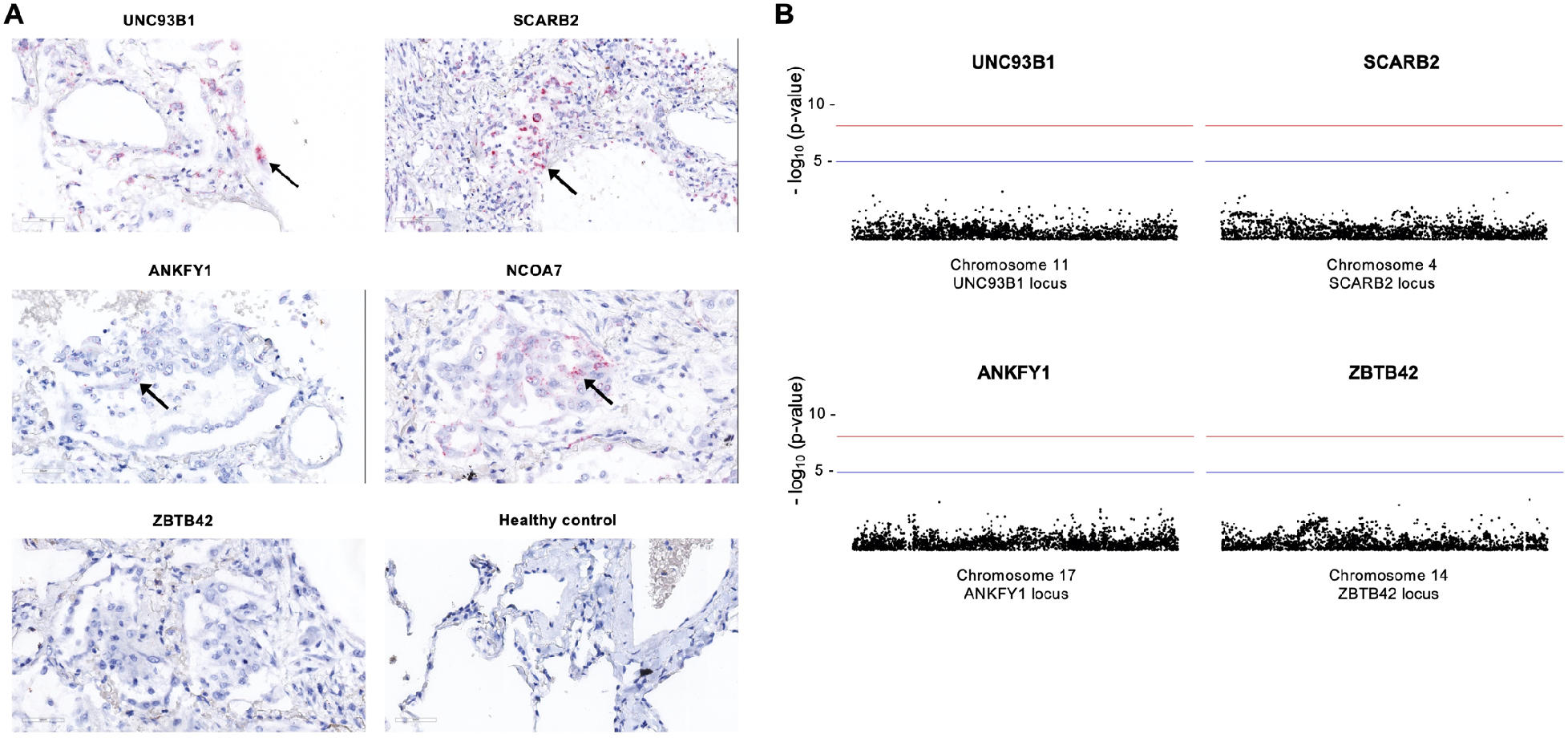
The ISG OAS1 strongly inhibits SARS-CoV-2. (A) Detection of UNC93B1, SCARB2, ANKFY1, NCOA7, ZBTB42 gene expression by RNAscope in FFPE lung tissue of deceased COVID-19 patients compared to healthy control lung tissue. No signal detected for ZBTB42 while gene expression could be detected for the other genes. Arrows indicate staining +ve cells. (B) Metaanalysis of the COVID-19 Host Genetics Initiative (covid 19hg.org) for genetic variation between critical ill COVID-19 patients and control populations at the gene locus of the UNC93B1, SCARB2, ANKFY1 and ZBTB42 genes with the red line indicating the threshold for significant SNPs (yellow dots).

**Supplementary Figure 4.**
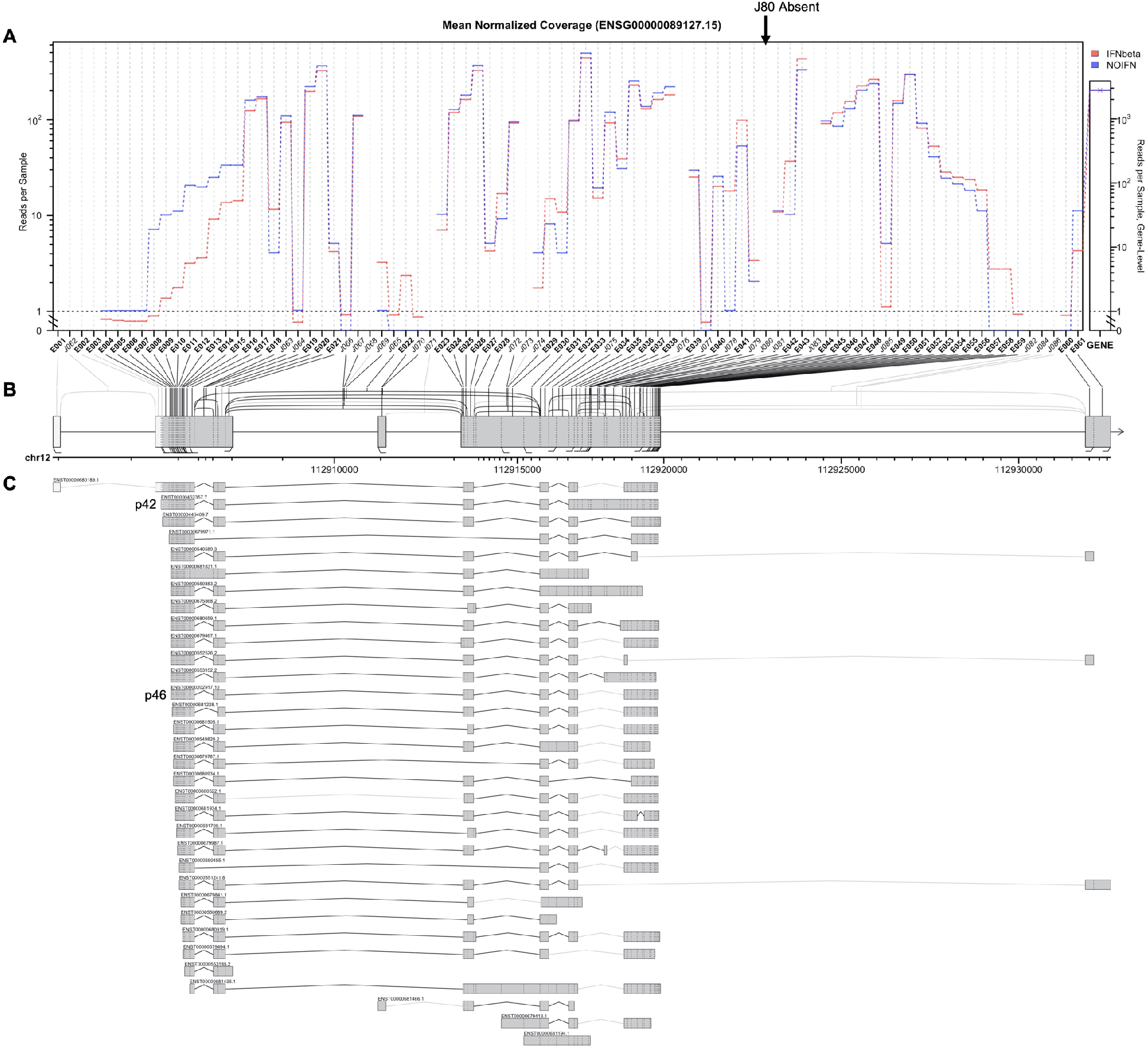
Transcripts encoding P46 were not detected using JunctionSeq differential usage analysis of OAS1 transcripts from AA homozygous A549 cells. (A) The expression levels at each junction and exon for every annotated OAS1 transcript (Ensembl) from A549 cells stimulated with IFN (red) or not treated (blue). Junction 80 (380) is highlighted with an arrow. (B) A diagrammatic representation of introns and exons on chromosome 12, highlighting the corresponding chromosomal location to the transcript junctions. (C) A diagrammatic representation of the differential exon usage of all known OAS1 transcripts (Ensembl). Splice junctions detected in the RNAseq data are linked by black lines whereas undetected junctions are linked by light gray lines. The transcripts encoding p46 and p42 proteins are highlighted.

**Supplementary Figure 5.**
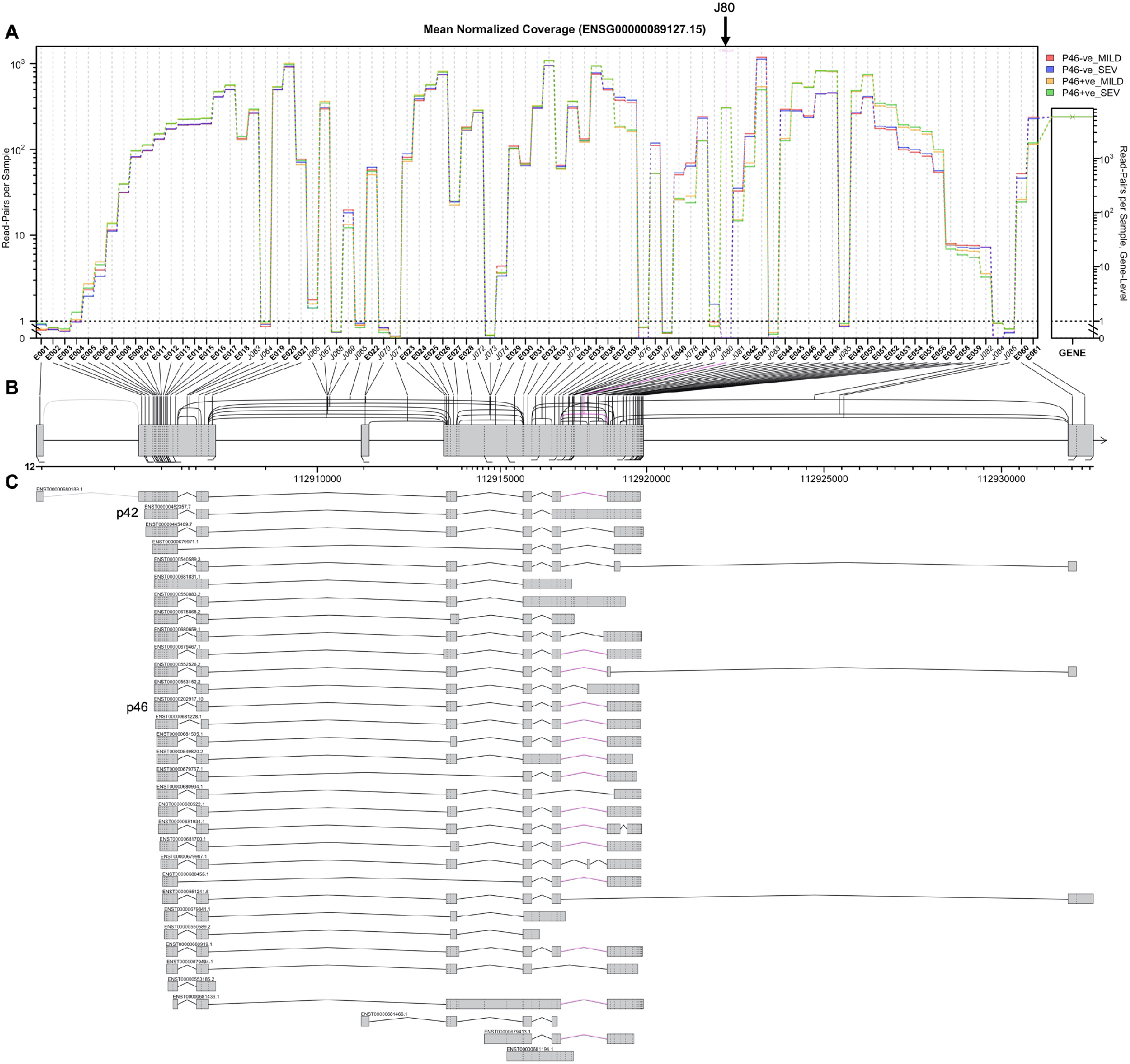
Strand-specific exon and known splice-junction loci counts of OAS1 transcripts generated by QoRTs from 499 samples were used to stratify patients into p46 transcript positive and negative patients and subjected to differential usage analysis via JunctionSeq. (A) The expression level at each junction and exon for every annotated OAS1 transcript (Ensembl) from 499 COVID-19 patients. The mild and severe patient groups were subdivided based on presence or absence of junction 80. Patients with no mapped reads were classified as p46 -ve and patients with mapped reads as p46 +ve. Junction 80 (180) is highlighted with an arrow. (B) A diagrammatic representation of introns and exons on chromosome 12, highlighting the corresponding chromosomal location to the transcript junctions. (C) A diagrammatic representation of the differential exon usage of all known OAS1 transcripts (Ensembl). Splice junctions detected in the RNA-seq data are linked by black lines whereas absent junctions are linked by light gray lines. The transcripts encoding p46 and p42 proteins are highlighted. Junctions significantly different between groups are highlighted in purple.

## Notes

### Competing Interest Statement

The authors have declared no competing interest.

### Author Declarations

Ethical approval was given by the South Central-Oxford C Research Ethics Committee in England (reference 13/SC/0149), and by the Scotland A Research Ethics Committee (reference 20/SS/0028). The study was registered at https://www.isrctn.com/ISRCTN66726260. This work (ethics approval number 32077020.6.0000.0005) was approved on May 2020 by the National Committee in Ethics and Research, Brazil. in COMISSAO NACIONAL DE ETICA EM PESQUISA.

